# Analysis of national and international guidelines on respiratory protection equipment for COVID-19 in healthcare settings

**DOI:** 10.1101/2020.04.23.20077230

**Authors:** Gabriel Birgand, Nico T. Mutters, Jonathan Otter, Vanessa M. Eichel, Didier Lepelletier, Daniel J. Morgan, Jean-Christophe Lucet

## Abstract

**Background:** Consistent guidelines on respiratory protection for healthcare professionals combined with improved global supply chains are critical to prevent COVID-19. We analysed the guidelines published by national and international societies/organizations on facemasks and respirators to prevent COVID-19 in healthcare settings.

**Methods:** From the 1^st^ January to the 2^nd^ April 2020, guidelines published in four countries (France, Germany, United States, United Kingdom), and two international organizations (US and European Centre for Diseases Control, and World Health Organization) were reviewed to analyse the mask and respirators recommended for healthcare settings during the COVID-19 outbreak. The aerosol generating procedures (AGP) definitions and the strategy recommended for optimizing supplies and overcoming shortages were collected.

**Findings:** The recommendation of respirator was universally recommended for AGP across countries, although the type of respirators and what constituted an AGP was variable. Some guidance maintained the use of N95/99 for all contact with confirmed COVID-19 cases (i.e. Germany) whereas others, recommended a surgical mask (i.e. WHO, UK, France). Most guidelines were published in March with either downgraded (US and European CDC), relatively stable (WHO, Germany, and UK), or a mixing of high and low level equipment (France). The strategies to overcome shortage of respiratory protection equipment were based on minimizing the need and rationalizing the use, but also prolonging their use, reusing them after cleaning/sterilization, or using cloth masks.

**Interpretations:** In a crisis context, stable and consistent guidelines clearly detailing the respiratory protection type, and their indications, may prevent the confusion and anxiety among frontline staff, and avoid shortage.

## INTRODUCTION

The coronavirus disease 2019 (COVID-19) pandemic surprised many with the rapidity of spread. Healthcare systems from many countries have become overwhelmed by infected or infectious patients requiring care and respiratory support. ^1^

The effective use of personal protective equipment (PPE) including facemasks or air-purifying respirators is critical to protect the wearer from the transmission of droplet and airborne pathogens.^2^ The evidence is limited for the assessment of the PPE requirements for the prevention of transmission of SARS-CoV-2 virus that causes COVID-19. The transmission mode of SARS-CoV-2, initially unclear, led to the implementation of high-level precautions, including airborne and contact, to prevent the spread to and from HCP and patients. The increasing knowledge on SARS-CoV-2 in addition to the SARS-CoV-1 background, suggested droplets and hand contaminations as the main transmission route. ^3^ In an open room, putting beds at a 1·5 meters distance was described to be strongly associated to the absence of transmission of SARS-CoV-1. ^4,5^ However, procedures on the respiratory tract may generate the aerosolization of small particles, with potential airborne transmission. ^6,7^ Moreover, the transmission from asymptomatic and presymptomatic persons, not recognized as having SARS-CoV-2 infection and therefore not isolated, might contribute to further spread. ^8^ This emerging understanding of the SARS-CoV-2 transmission dynamics combined with a global shortage of facemasks and respirators, led to recommendations for a lower-level of respiratory protection.^3^ Many recommendations has been published worldwide following the emergence of COVID-19, based on variable evidence. Consistent guidelines on PPE based on rational data and evaluation of risk, combined with improved global supply chains for the necessary PPE are critical to protect staff and patients.

We present a narrative review with critical summary and comparison of the latest guidelines published by national and international societies/organizations on facemasks and respirators to prevent the spread of SARS-CoV-2 in healthcare settings. We describe the content, and analysed the number of recommendations, the timeline according to the epidemic evolution, the consistency of guidelines over time, and the strategies to optimize supplies and overcome shortages.

## METHODS

From the 1^st^ January to the 2^nd^ April 2020, guidelines published in four countries (France, Germany, United States, United Kingdom), and two international organizations (European and US Centre for Diseases Control, and World Health Organization) were reviewed to analyse the mask and respirators recommended as PPE for the care of patients with COVID-19. The review and data extraction were performed by infection control specialists involved in COVID-19 prevention and control in each country. Guidelines published by national societies of infection control, Infectious diseases, intensive care, emergency medicine, anaesthesia, surgery, occupational medicine, endoscopy, neonatology and obstetric, nursing, public health authorities or ministries were conveniently considered. The identify of additional guidelines was performed by snowball sampling. Guidelines were eligible for analysis if they (1) included specific guidelines, (2) were written for HCP protection, (3) targeting healthcare settings. For each included guideline, the following information were extracted: the organization or society publishing the guideline, the date of publication, the type of facemask/respirator recommended, stratified by the circumstance/indication of use, and lists of aerosol generating procedures (AGP) (when present). The strategy recommended for optimizing supplies and overcoming shortages was collected (if included).

## RESULTS

At the international level, the WHO issued four guidelines during the study period, consistently recommending the use of “medical” masks for direct contact with COVID-19 patients, and respirator FFP2 (N95) for AGP on those patients.^9–11^ (Table 1) The ECDC published two technical reports, with two updates for the second report. The recommendations moved from FFP2 or FFP3 (N99) respirators when contact with suspected or confirmed COVID-19 patients early February, to switch to the use of surgical mask (SM) in absence of respirators in the following updates.^12,13^ (Figure 1)

**Table 1.**
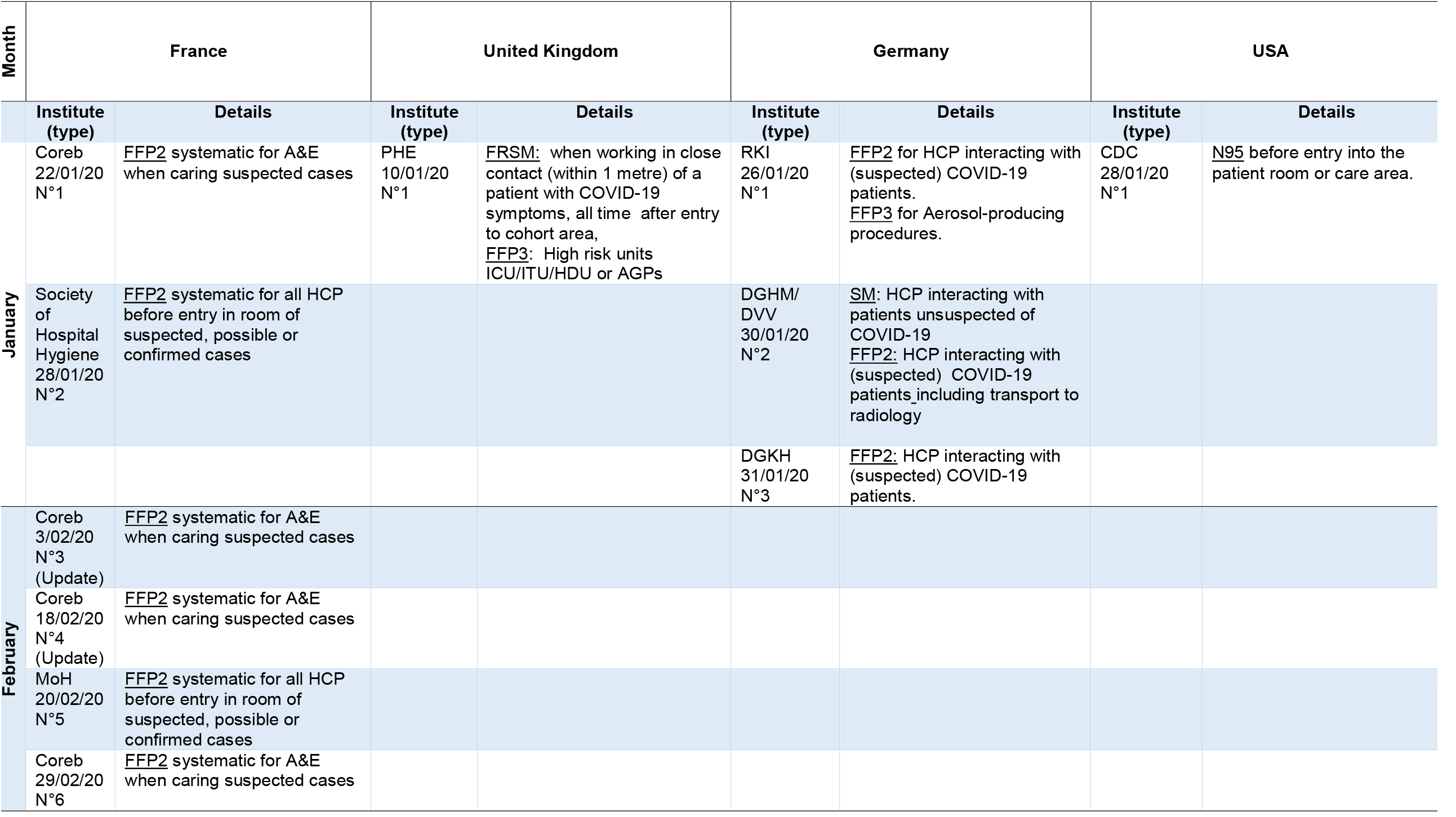
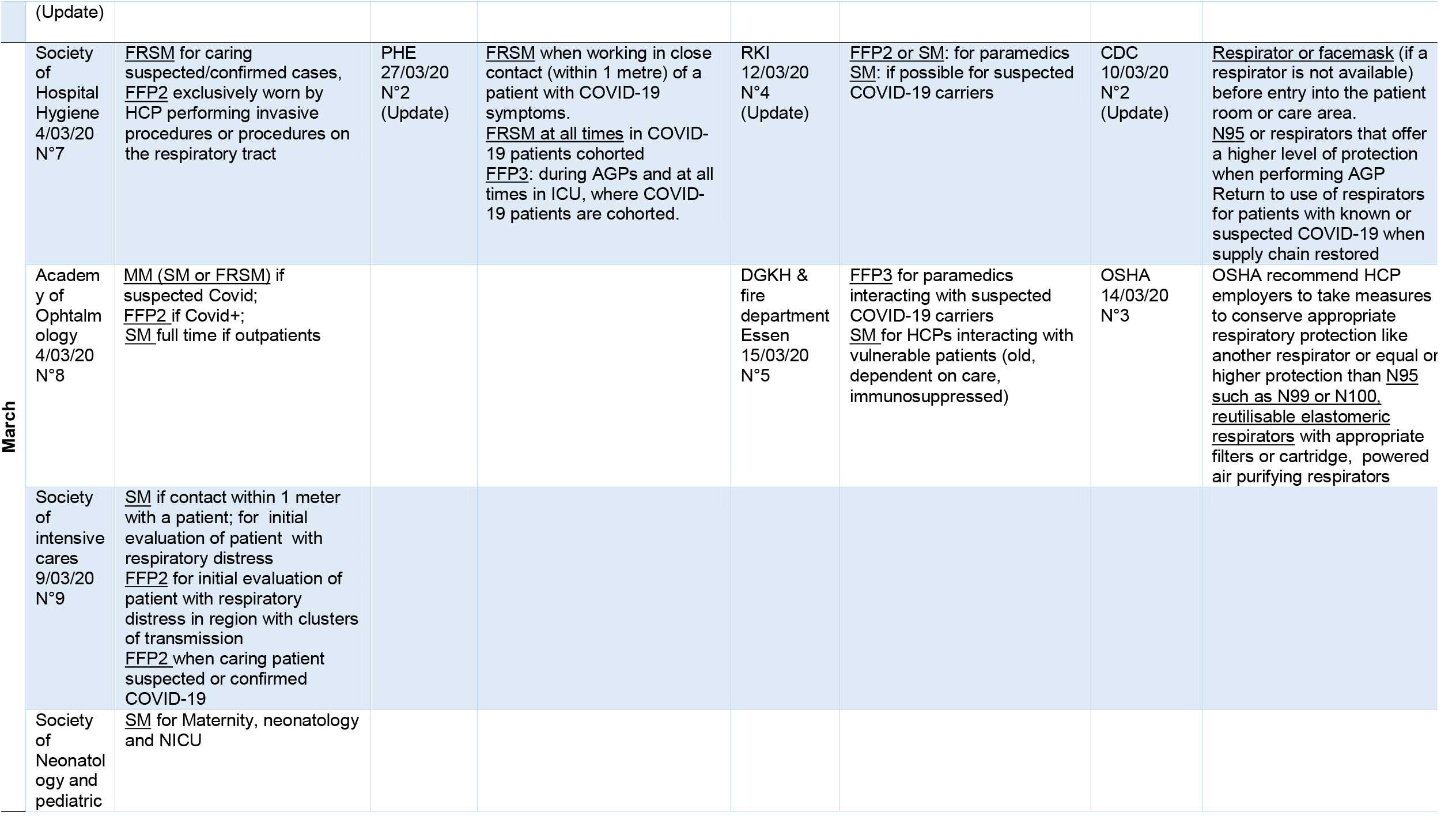
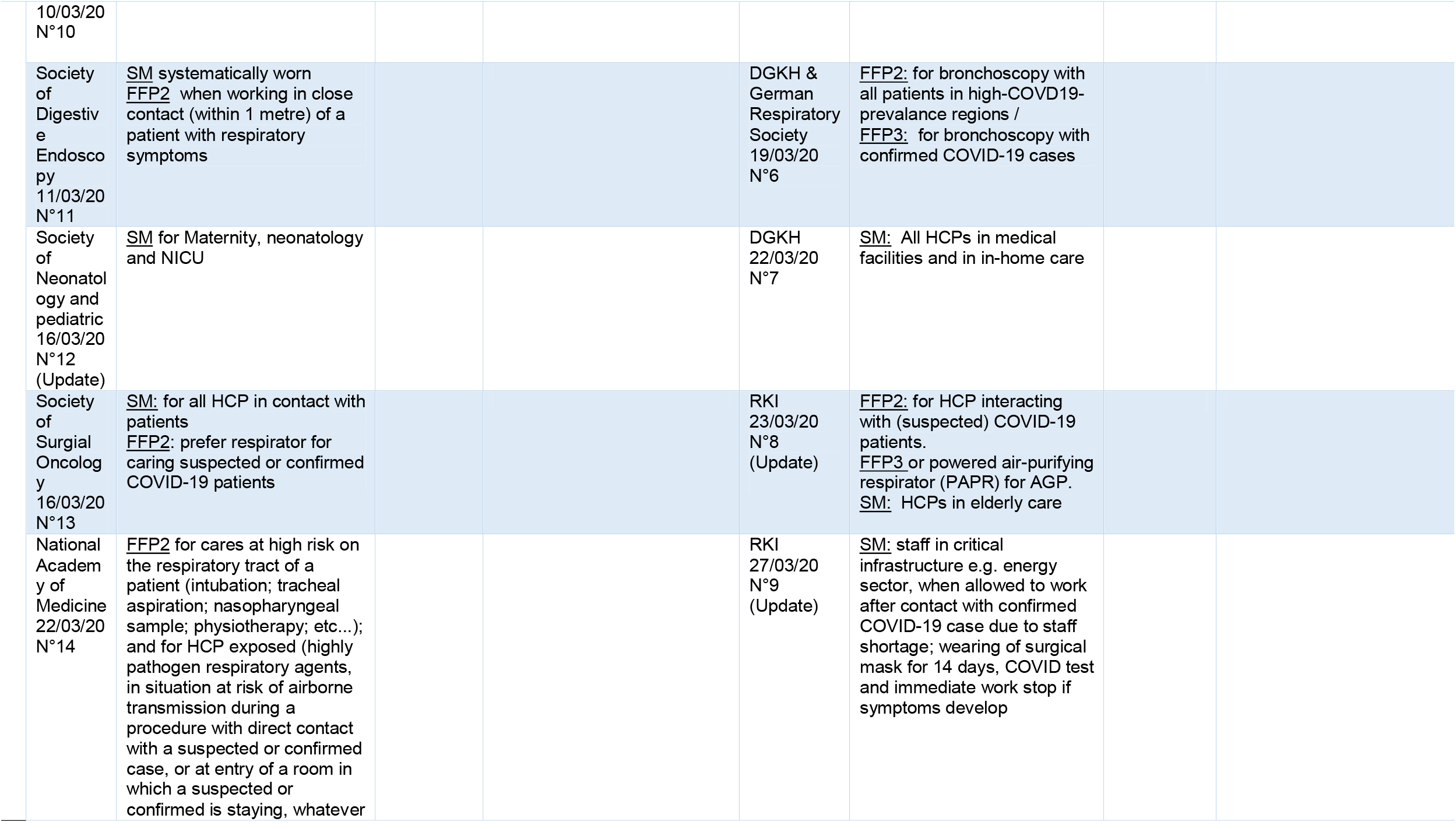
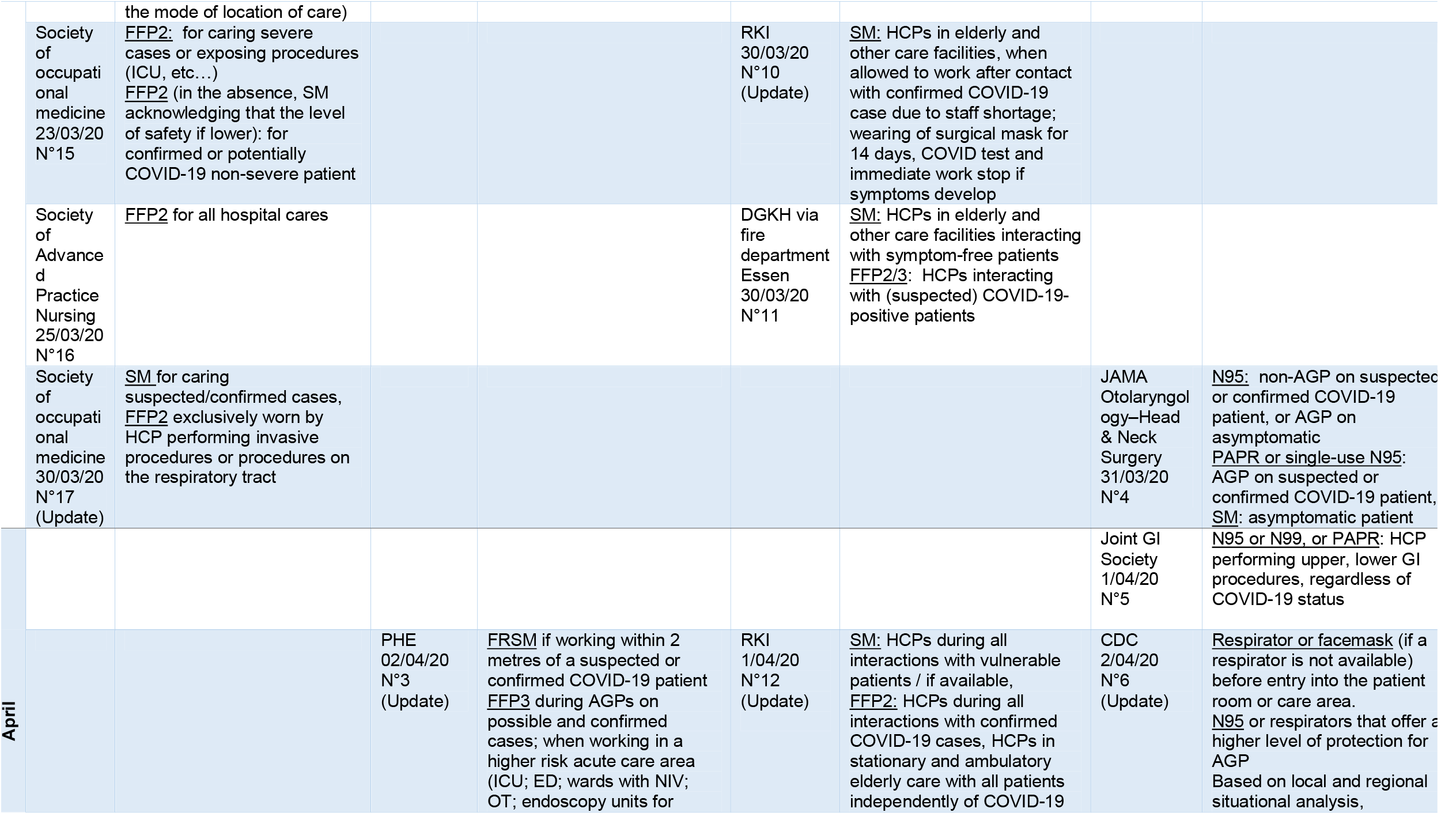
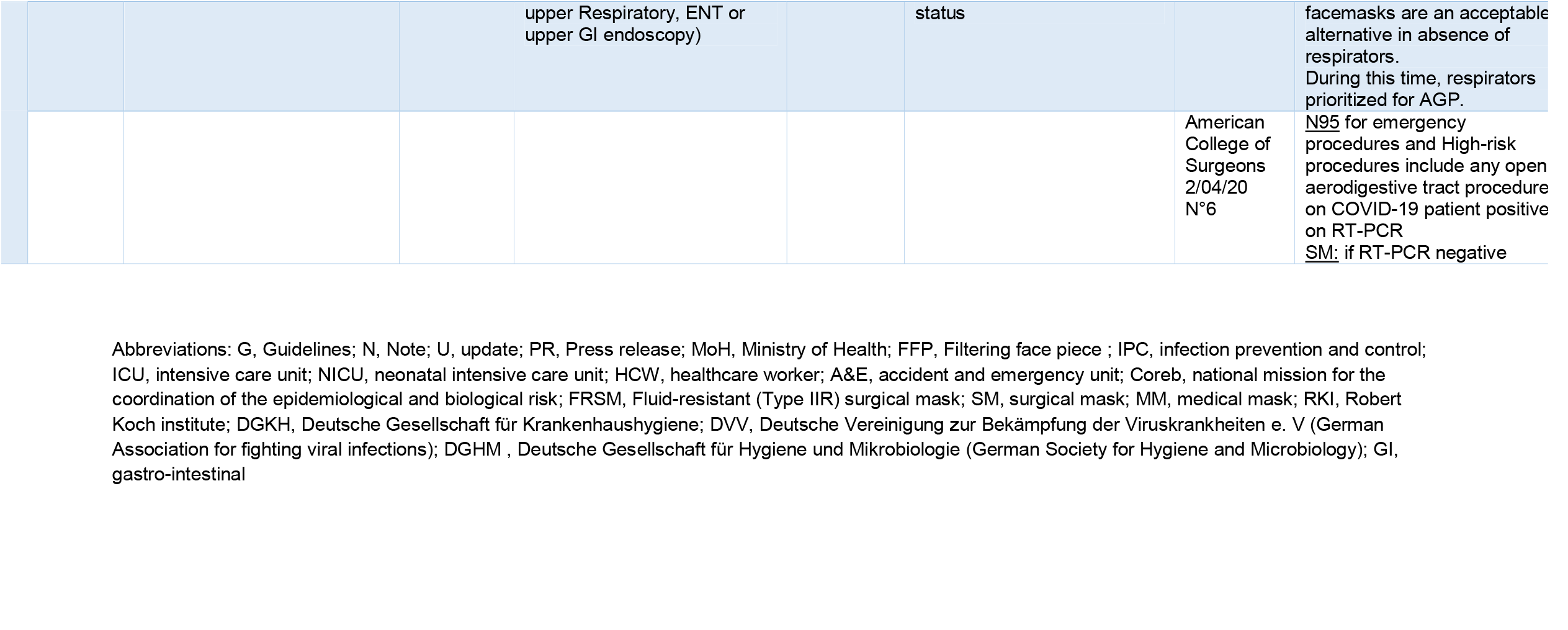
Description of the national and international guidelines published on type of facemask and respirator recommended in the COVID-19 context, according to circumstances, in France, England, Germany and USA.

**Figure 1:**
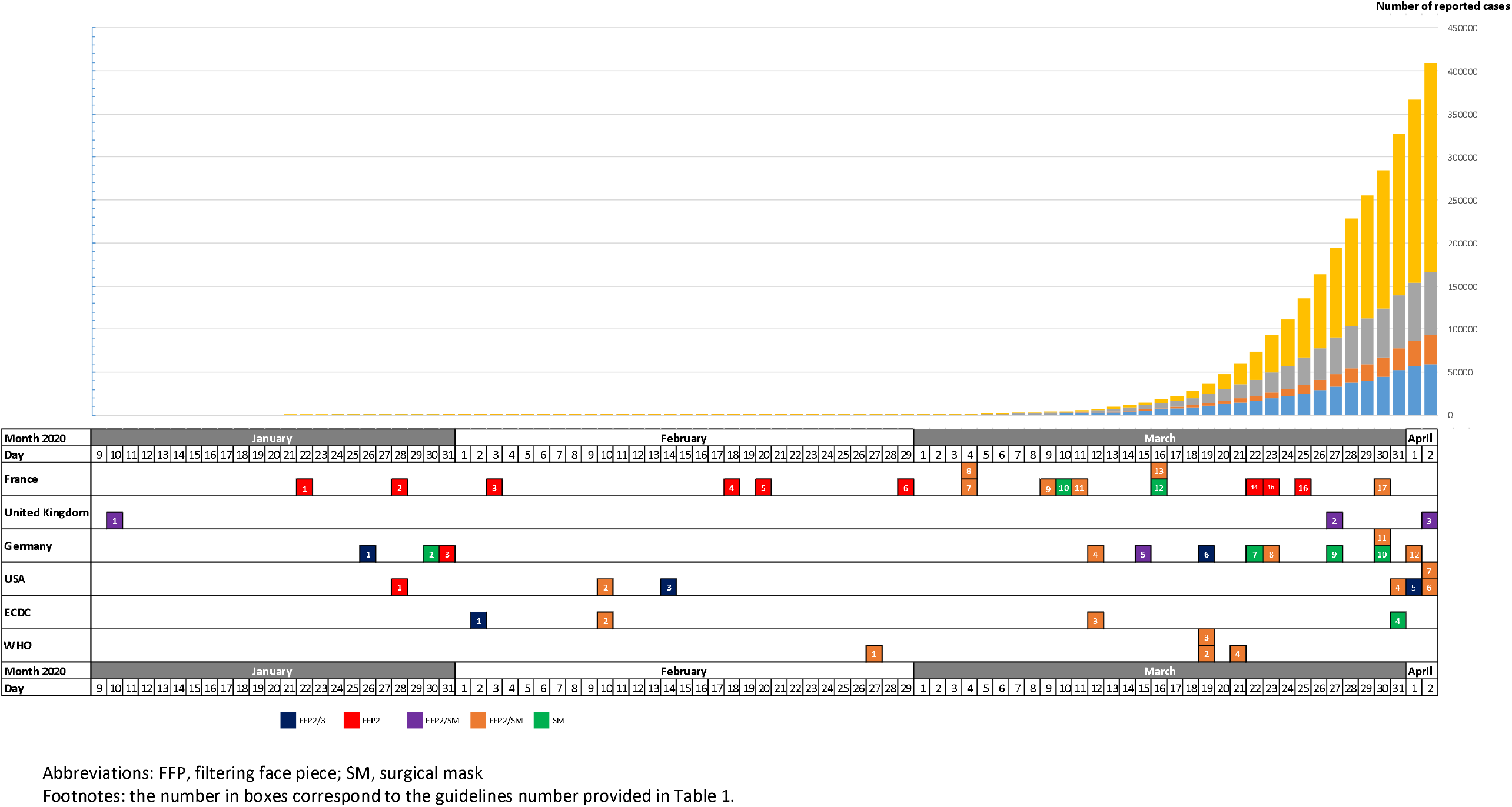
Chronological evolution of guidelines according to the national epidemiology of COVID-19.

In the US, 19 societies, colleges or organizations webpages were checked for COVID-19 guidelines. Five guidelines were published detailing respiratory protections for HCP caring patients with suspected or confirmed COVID-19, including one from the CDC updated twice.^14^ Initially, CDC recommended at least the use of N95 respirator before entry into the patient room or care area. Then, the guidance changed by extending the possibility to use N95 respirators or respirators that offer a higher level of protection used when performing or present for an AGP. The Occupational Safety and Health Act (OSHA) recommended HCP employers to take measures to conserve appropriate respiratory protection like another respirator or equal or higher protection than N95 filtering facepiece such as N99 or N100, reusable elastomeric respirators with appropriate filters or cartridge, powered air purifying respirators.^15^ Otolaryngology–Head & Neck Surgery specialists published their recommendations in a scientific journal.^16^ The N95 respirators was advised for non-AGP on COVID-19 patient, or AGP on asymptomatic, whereas a powered air purifying respirators (PAPR) was added for AGP COVID-19 patient. A joint guidance from gastro-intestinal (GI) societies recommended N95 or N99, or PAPR for HCP performing upper, lower GI procedures, regardless of COVID-19 status.^17^ The latest CDC guidelines introduced the notion of facemasks being acceptable alternative when the supply chain of respirators cannot meet the demand based on local and regional situational analysis. In period of shortage, available respirators were to prioritize for procedures that are likely to generate respiratory aerosols. Finally, the American College of Surgeons recommended N95 for emergency procedures and high-risk procedures include any open aerodigestive tract procedure on COVID-19 patient positive on RT-PCR, and SM if RT-PCR negative.^18^

A total of 15 different societies and organizations were checked for COVID-19 guidelines in the UK. All were referring to the UK government webpage for PPE recommendations. A unique guideline was issued by Public Health England (PHE), and updated twice.^19^ The first version recommended fluid-Resistant SM when working in close contact (within one meter) of a patient with COVID-19 symptoms, all time after entry to cohort area, and FFP3 when working in high risk units (i.e. ICU) or AGPs. Early April, the recommendations were changed with SM for any clinician working in a hospital, primary care or community care setting within two meters of a suspected or confirmed coronavirus COVID-19 patient. The latest guidance recommends sessional use (for more than one patient) of an SM when working in clinical areas with confirmed or suspected COVID-19 cases, even when two meters away from patients. The FFP3 was recommended during AGPs on possible and confirmed cases, regardless of the clinical setting, and when working in a higher risk acute care area (i.e. intensive cares). SM are also recommended when delivering care to vulnerable patients who are undergoing shielding. The guidance also goes further for clinical settings in which there is sustained transmission. In these settings, an FRSM should be worn for all patient contact (regardless of COVID-19 status) and an FFP3 respirator for all AGPs (regardless of COVID-19 status).

In Germany, the first guideline was issued by the Robert Koch Institute (RKI), and recommended the use of N95 respirators for HCP interacting with (suspected) COVID-19 patients, and FFP3 for AGPs. This version was followed by five updates.^20^ The systematic worn of SM was recommended for HCPs suspected of COVID-19, and those working in elderly care, or critical infrastructure. The latest guideline from RKI advised a FFP2 for HCPs during all interactions with confirmed COVID-19 cases. The German Society for Hygiene and Microbiology (DGHM) & German Association for fighting viral infections (DVV) early recommended the systematic worn of SM for HCP interacting with patients unsuspected of COVID-19, and FFP2 if suspected COVID-19 patients including transport to radiology.^21^ The Deutsche Gesellschaft für Krankenhaushygiene e.V. (DGKH; German Society of Hospital Hygiene) issued five documents, mainly recommending the FFP2/3 for HCPs interacting with, or performing AGP on suspected or confirmed COVID-19 patients. The SM was advised for HCPs interacting with vulnerable patients.^22^

In France, 12 different guidelines from 11 different societies/organizations (two from the French Society of Hospital Hygiene) were published within the three-month period.^12,23–32^ Three guidelines were updated from one to three times. An acceleration in publication and discrepancies across guidelines were observed in March. Globally, guidelines started with the use of FFP2 respirators in all circumstances, with a switch to surgical mask for direct care cares and FFP2 for AGP at the growing phase of the outbreak mid-march. Some societies published recommendations on the systematic worn of SM if contact with patients, including non-infected. A mix of high and low level measures were observed at the end of the studied period, with some guidelines recommending the FFP2, and others SM for all cares, or for confirmed or potentially COVID-19 non-severe patient at the end of the studied period.

### Aerosol generating procedures (AGP)

The US CDC, ECDC and WHO provided few examples of AGP, mainly based on the cumulated experience for influenza, MERS-CoV and Ebola. The most detailed list was published by Public Health England in UK with nine different procedures. For France, the list was not published despite giving recommendation of FFP2 respirators for performing AGP. The only assessment of the evidence was performed by the Health Protection Scotland (HPS) which graded the PHE AGP list according to a literature review, with two categories: B for all procedures excepted the high flow nasal oxygen in C.^33^ Despite various lists according to countries and institutions, some core procedures were consistently cited, including the tracheotomy, bronchoscopy, non-invasive ventilation and induction of sputum. (Table 2) While dental procedures were mentioned in the national but not in international guidelines, surgery and post-mortem procedures, invasive ventilation and nasal oxygen were inconsistently listed. Several organizations in the US published their own AGP list. The American College of Surgeons considered as AGP any open aerodigestive tract procedure.^18^ The Otolaryngology–Head & Neck Surgery specialists included the endoscopy on the GI and respiratory tract, powered instrumentation in mucosal head and neck surgery, possibly laparoscopic surgery, drainage of peritonsillar abscess, placement of devices in the nose or airway.^16^ The American Gastroenterological Association listed the upper GI endoscopic procedures such as esophagogastroduodenoscopy, small bowel enteroscopy, endoscopic ultrasound, endoscopic retrograde cholangiopancreatography (ERCP), breath tests, and esophageal manometry. ^34^ Finally, the Society for Interventional Radiology detailed a long list of invasive procedure including biopsies, drains, tube placement, and any procedure that may induce coughing.^35^

**Table 2:**
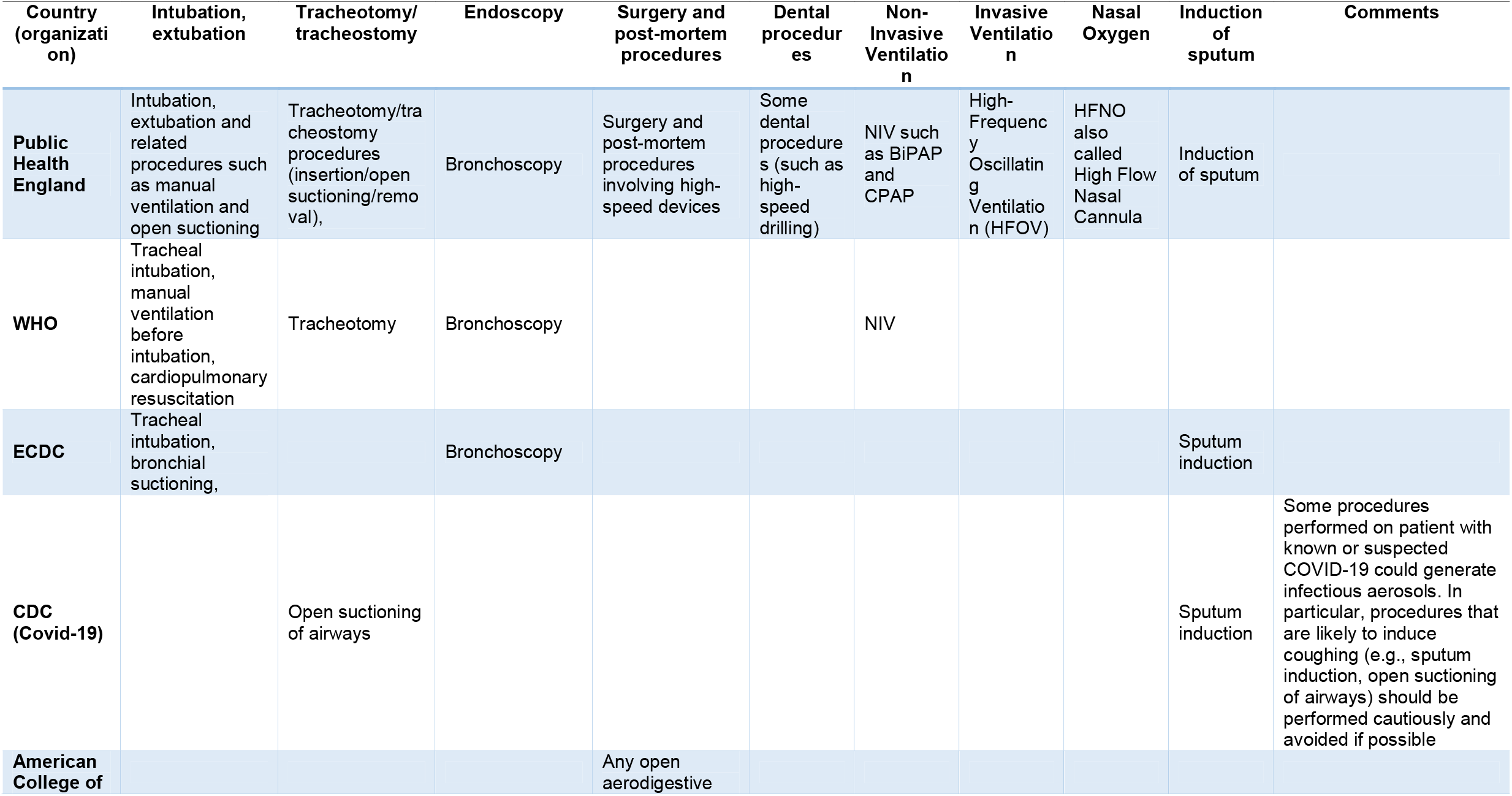
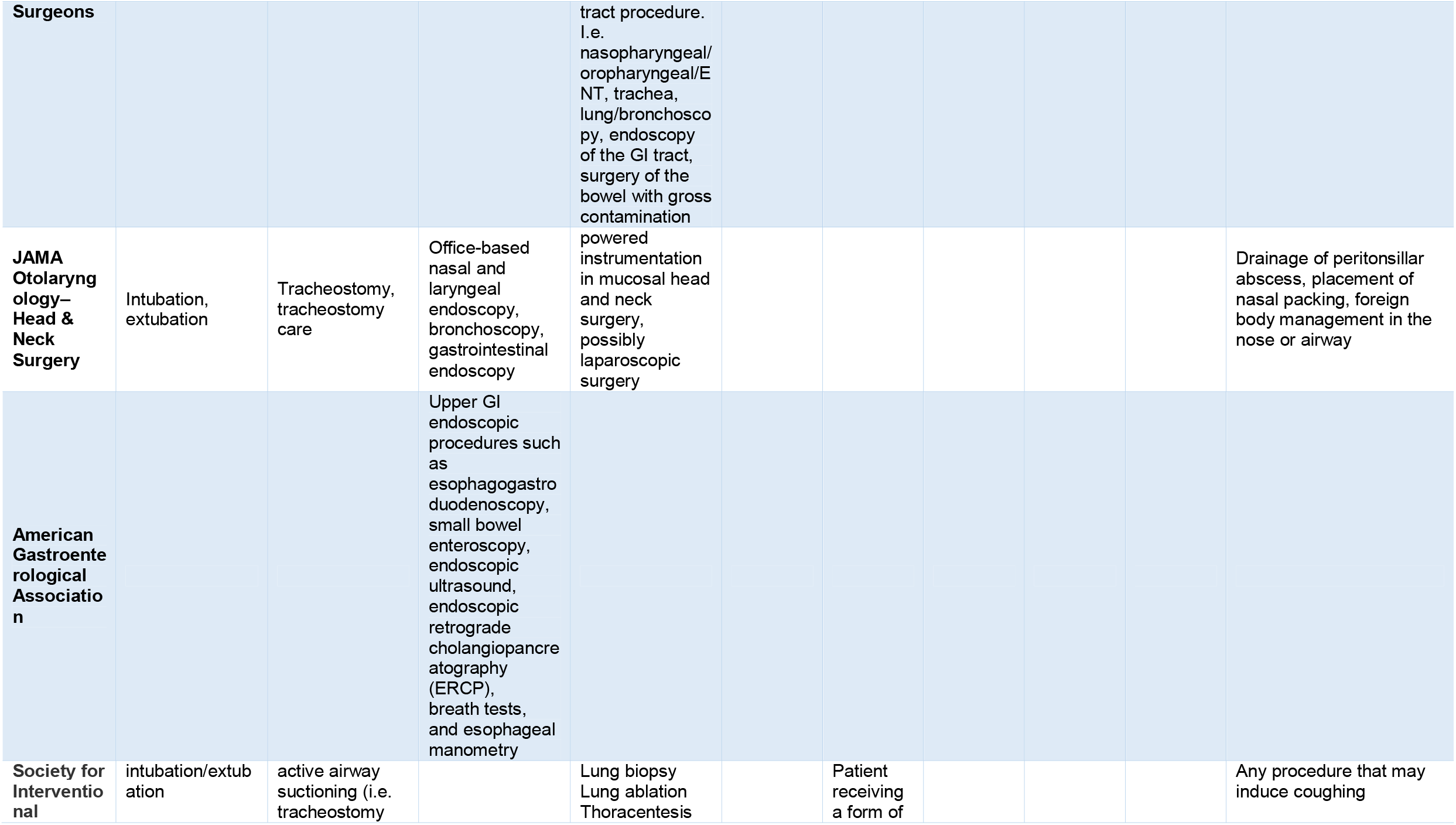
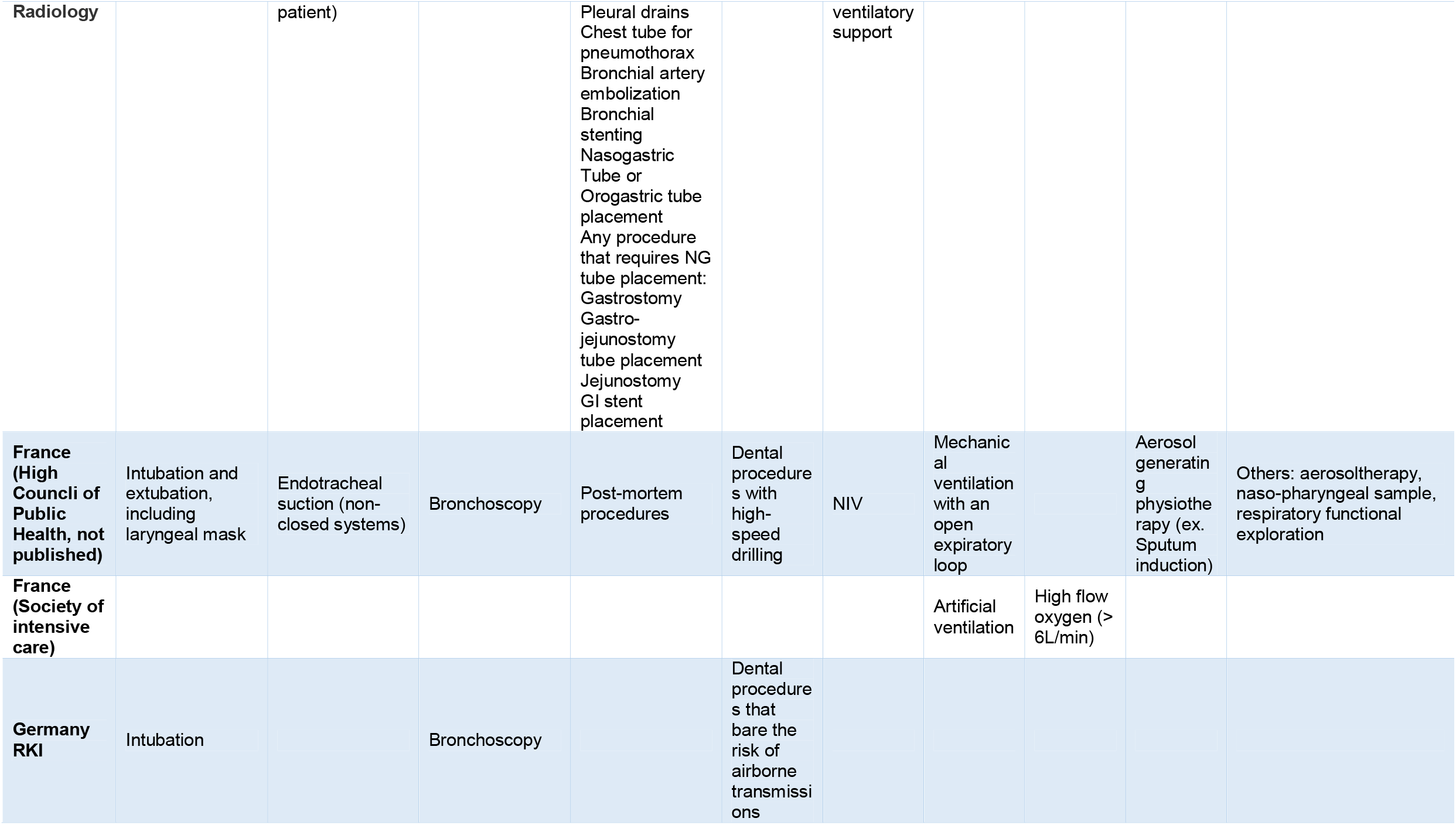
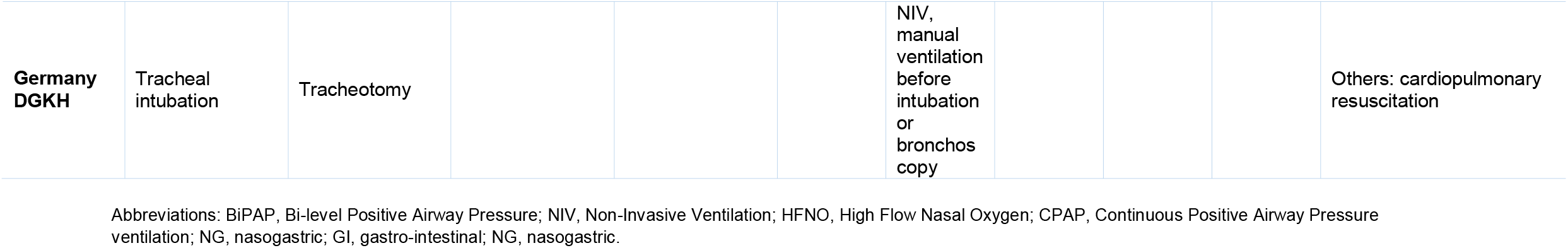
Description of the aerosol generating procedures listed in national and international guidelines.

### Strategies to optimize supplies, and overcome shortage

The first strategy employed to optimize supplies was based on prolonging the use of respiratory protection equipment. The ECDC allowed the use of respiratory protection equipment up to 4-6 hours for respirators when not removed or damaged. The WHO allowed wearing the same N95/99 respirator no more than four hours, whereas France extended the duration of use of SM with a maximum duration of four hours, and eight hours for a FFP2 respirator. PHE advised to wear a SM or FFP3 at all times rather than only when in close contact with a patient, with sessional use (defined as a period of time where a HCP is undertaking duties in a specific care setting/exposure environment) in inpatient areas, and single use (disposal after each patient and/or following completion of a procedure, task, or session) or sessional use in operating room, labour area, or for patient transfers.^19^ The PHE argued this position by the absence of evidence on the risk reduction of infection transmission, due to frequent handling of this equipment when changing between patients visits. The US CDC early stated the possibility to use N95s exceeding shelf life. The inspection of the visual integrity of any part of the respirator were recommended if reutilization.

The reuse of masks was recommended in several countries. For ECDC, the reuse of single-use respirators (FFP2 and FFP3) after cleaning/sterilization was considered as a last-resort method. In Germany, in period of shortage of supplies, the re-use of personalized FFP2/3was made possible if only worn by one person / decontamination of masks (30min 65-70°C) is possible for CE-certified masks and other masks that were (quickly) self-tested for heat stability. CDC and NIOSH did not recommend that F N95/99 be decontaminated or reused as standard care. However, in times of crisis, this option may need to be considered when FFP2/3 shortages exist, recommending ultraviolet germicidal irradiation (UVGI), vaporous hydrogen peroxide (VHP), and moist heat showing the most promise potential for decontamination. In absence of manufacturer or third-party guidance or procedures, these FFP2/3 should not be worn by HCPs when performing or present for an AGP.

The ECDC proposed the use of the surgical mask, and home-made cloth masks when a shortage was severe. Facing critical shortage, the French Society for Infection Control opened the possibility for using facemasks made in SMS sterilization wrap for non-frontline staff based on the evaluation of filtration capacities of the material. However, the use of other types of facemasks (i.e. Cloth masks, paper masks, towel…), was banned in healthcare settings due to lacking scientific data on the efficacy. In Germany, some guidelines also established a hierarchy, with first the possibility to replace FFP3 by FFP2 by surgical mask according to availability, and secondly to replace the FFP3 by FFP2 by surgical mask by “community” textile mask according to availability. For the US CDC, when no facemasks are available, the strategy rely on HCP immunity or severity risks, the use of physical barriers or distancing (face shield). The use of homemade masks (e.g., bandana, scarf) for care of patients with COVID-19 is considered as a last resort.

The WHO strategy focused on minimizing the need (use other barriers, restricting entries in the rooms of COVID-19 patients), rationalizing the use (strict indications, adapted to setting, type of HCP and activity). In addition, a centralization and coordination of the supply chain is recommended. The CDC categorized three strategies according to situations: the conventional (face masks are used in conventional ways), the contingency (avoiding the extra use of masks and replace respirators by the use of facemasks), and the crisis capacity strategies (extension beyond manufacturer shelf life and the re-use of facemasks).

## DISCUSSION

In this review of national and international guidelines, the recommendation of FFP2/3 for AGP was the universal measure across countries, although what constituted an AGP was variable. Some guidance maintained the use of FFP2/3 for all contact with confirmed COVID-19 cases (i.e. Germany) whereas others, recommended a surgical mask (i.e. WHO, UK, France). The WHO, since the first guide publication, recommended the “medical” surgical mask for HCPs providing direct care to COVID-19 patients, and respirators for AGP. The ECDC and the US CDC downgraded the measures starting with N95/99 for all situations, to the consideration of mask with the highest available filter level, such as a surgical mask if N95/99 not available. In comparison, the national guideline published by China CDC recommended the SM for all staff in all healthcare facilities, the N95 in COVID patients area, and the N95 or PAPR when directly in contact confirmed cases, or during AGPs.^36^

The publication process varied according to countries and organization. The WHO guidelines appeared as the most consistent over time, with four publications keeping the same line on respiratory protection in healthcare settings. In the UK, only one guidance was released, and updated according to the time. This publication strategy avoids inconsistencies, providing a strong and harmonized practices across UK countries. On the other hand, 12 guidelines were published in France by 11 different societies or organizations, with major inconsistencies. In a similar but less intense manner, 12 recommendations were published by four different German organizations, of which two were published as a cooperation of two entities each. Some vagueness on the type of protection to use (the terms “medical”, “surgical” or “fluid resistant” facemask), and their indication (definition of high-risk units, AGP) may led to confusion from frontline staff.

Masks may play a symbolic role helping HCP in their daily duty by giving a sense of safety, well-being, and trust in their hospitals. The anxiety from HCP, emphasized by the mass communication in the press and social media, generate an irrational by understandable behaviours leading to the use a full panoply of protection equipment. In a period of confusion and sometime panic, the stability and precision in recommendation appear as a strength, for a better understanding by frontline staff, and ease the education from infection control specialists. The confusion around PPE guidance not rooted by science but rather by possible PPE shortage, leads to inappropriate use, and to potential shortages, with more exposed HCP as a consequence. The recommendation of N95/FFP2 as first line with surgical masks being considered acceptable in ECDC and CDC guidelines created a pressure to use N95/FFP2 inordinately early in the outbreak for rule-out cases with the unintended consequence of many facilities in the US needing to move to surgical mask only outside of AGP earlier for COVID-10 positive patients. This contributed to staff to believing they received inadequate protection only due to lack of supply. The tunnel vision on mask and PPE in general deviates the awareness from the compliance with standard measures such as hand hygiene. However, a large part of the individual contamination and cross transmission from patient to patient may occur through hands. The use of PPE and especially facemasks and respirators must be associated with a strict compliance with hand hygiene.^37^

Overall, the type of masks cited in guidelines was heterogeneous and sometimes confusing. Respiratory protections can be categorized in respirators and facemasks. Among respirators, some organizations recommended the use of FFP3, some FFP2 respirators, and others both according to procedures. These disposable respirators, are subject to standards specifying certain required physical properties and performance characteristics. One notable comparison point is the flow rates for the inhalation and exhalation resistance tests, and the filter performance stopping at least 95% for N95 and 99% for N99. Furthermore, in Germany, official institutions use the term “respirator” for powered air-purifying respirator (PAPR) which also leads to confusion in the international context. Among facemasks, the terms of medical, surgical and fluid-resistant facemasks were used, sometime without clear description. However, the facemask category relies on a filtration efficiency level (type I with ≥ 95%, and type II ≥ 98%), and a splash fluid resistance capacity (> 120 Hg mm). A clear and detailed type of facemasks may ease the interpretation of current national and international guidelines to prevent COVID-19.

AGPs are commonly defined as any medical and patient care procedure that results in the production of airborne particles (aerosols) 5 micrometres (μm) in size which can remain suspended in the air, travel over a distance and may cause infection if they are inhaled. ^38^ The actual knowledge on AGP, whether they confer a higher risk of transmission of infectious agents is scarce. In this absence of strong evidence, precautionary approaches were taken by most all countries of the present evaluation. The same issue was observed during the H1N1 pandemic, MERS and Ebola with less intensity as the phenomenon did not become pandemic and not affected high-income countries. The most recent assessment by WHO (2014) stated that there is only consistent evidence that there is an increased risk of transmission for the following procedures: tracheal intubation, tracheotomy procedure, non-invasive ventilation, and manual ventilation before intubation as AGPs.^38^ This evaluation was based on a systematic review of extremely limited volume and quality of studies requiring caution interpretation. ^39^ The current lists of procedures that is provided in guidance may have major logistic and psychological impact when trying to implement on the front line. Establishing evidence around AGP, their definitions, and defining a clear list would help better protect HCPs in their daily practice. The issue of variable AGP lists is how it tremendously impacts supply. A restrictive sense of AGP is including intubation, extubation, CPR and a few other procedures. On the other hand, a broad view includes any surgical procedures, with some guidelines requiring PAPRs in a negative pressure operating room, which is very burdensome and not possible in many hospitals. A lack of a standard CDC list meant the broad definition ruled in the US with potential supplies extra-use. Despite a narrow view of what an AGP would be helpful, it would also be contentious, probably explaining the vague definitions in most current guidelines.

One minor drawback of our study is, that despite the greatest possible care, it is possible that some updates of recommendations might have been wrongly dated. Some publishing entities overwrote their recommendation with an updated version and removed the older versions online without providing linked and dated previous documents.

Among national and international guidelines analysed, the N95/99 respirators was universally recommended when performing AGP. However, what constituted an AGP was variable. Variations existed for the respiratory protections type for HCP working in direct contact on suspected or confirmed COVID-19. Some countries relied on the FFP3/2 respirators and others on the surgical mask. Stable and consistent guidelines inside and across countries may prevent the confusion, for an appropriate use of masks, and avoid shortage. More scientific evidence is necessary for evaluating the risk of transmission of SARS-CoV-2 during AGP and to accurately use respiratory protections in healthcare settings.

## Data Availability

All data referred to in the manuscript are available online

## Declarations

### Ethics approval and consent to participate

not applicable

### Consent for publication

not applicable

### Availability of data and material

Data sharing not applicable to this article as no datasets were generated or analysed during the current study

## Acknowledgements

We thank Nina Zhu, Anne Savey, and Karine Blanckaert for their contribution to the review of guidelines.

